# Forecasting daily confirmed COVID-19 cases in Algeria using ARIMA models

**DOI:** 10.1101/2020.12.18.20248340

**Authors:** Messis Abdelaziz, Adjebli Ahmed, Ayeche Riad, Ghidouche Abderrezak, Ait-Ali Djida

## Abstract

Coronavirus disease has become a worldwide threat affecting almost every country in the world. The aim of this study is to identify the COVID-19 cases (positive, recovery and death) in Algeria using the Double Exponential Smoothing Method and an Autoregressive Integrated Moving Average (ARIMA) model for forecasting the COVID-19 cases.

The data for this study were obtained from March 21^st^, 2020 to November 26^th^, 2020. The daily Algerian COVID-19 confirmed cases were sourced from The Ministry of Health, Population and Hospital Reform of Algeria. Based on the results of PACF, ACF, and estimated parameters of the ARIMA model in the COVID-19 case in Algeria following the ARIMA model (0,1,1). Observed cases during the forecast period were accurately predicted and were placed within the prediction intervals generated by the fitted model. This study shows that ARIMA models with optimally selected covariates are useful tools for monitoring and predicting trends of COVID-19 cases in Algeria.

## 1. INTRODUCTION

On March 11^th^, 2020, the World Health Organization (WHO) declared COVID-19 as a worldwide pandemic. In December, 2019, a local outbreak of pneumonia of initially unknown cause was detected in Wuhan (Hubei, China), and was quickly determined to be caused by a novel coronavirus [1], namely severe acute respiratory syndrome coronavirus-2 (SARS-CoV-2) [2]. The outbreak has since spread to every province of mainland China firstly and has been propagated around countries and regions of the world. The contagious COVID-19 devastated normal life around the world. As of November 26^th^, 2020, COVID-19 has infected more than 60776978 confirmed cases in the world, has killed more than 1428228 people, and has forced more than 7 billion to stay in their homes [3]. In response to this ongoing public health emergency, an online interactive dashboard has been developed, hosted by the Center for Systems Science and Engineering (CSSE) at Johns Hopkins University, Baltimore, MD, USA, to visualize and track in real time reported cases of coronavirus disease 2019 (COVID-19) in the world [3].

Coronaviruses are a large family of viruses with some causing less-severe disease, such as the common cold, and others more severe disease such as MERS and SARS. Some transmit easily from person to person, while others do not.

According to Chinese authorities, the virus in question can cause severe illness in some patients and does not transmit readily between people.

The new coronaviruses lurking around the world are threatening our rules, and the prevalence of fear and panic is increasing. It has also affected the cryptocurrency market [4-5].

Algeria reported its first COVID-19 case, on February 25^th^, 2020. In November 26^th^, 2020, Algeria has reported 79110 confirmed cases with 51334 recoveries and 2352 deaths by COVID-19 [6].

Countries all over the world are challenged with this virus and have declared lockdowns in their various cities and states. The researchers estimate that the virus proliferates to more than two persons from every infected person, highlighting the possibility to infect millions [6]. In order to control this pandemic, Algerian government has instituted on March 23^rd^, 2020, several non-pharmaceutical intervention strategies. Thus strategies along with other measures such as social distancing, isolation and quarantine aimed to break the chain of transmission of COVID-19 in Algeria [7]. These measures were implemented with the aim to flatten the pandemic curve and prevent an exponential rise in new COVID-19 infections that would allow for the effective management and control of the pandemic.

Accurate forecasting of COVID-19 case trends is essential for the preparedness of health systems in terms of outbreak management and resource planning. Mathematical and statistical modeling of infectious disease is effective tools that would enable health systems to anticipate future disease trends [8-9]. Time series models such as the Autoregressive Integrated Moving Average (ARIMA) have been widely used to statistically model and forecast infectious disease trends [10]. ARIMA models are preferred in this context as they are suitable for investigations into short-term effects of acute infectious diseases and are a flexible class of models that are appropriate to fit several trajectories, and have been well documented in the literature [10-11]. ARIMA models have been used in several studies to forecast the COVID-19 outbreak trends [12-13-14-15]. However, to date, there are no studies conducted using ARIMA models to forecast the COVID-19 cases outbreak in Algeria. In this study, ARIMA models were developed using daily COVID-19 confirmed and active cases in Algeria to identify the best fitting model COVID-19 cases from March 21^st^, 2020 to November 26^th^, 2020. Forecasting future COVID-19 cases using ARIMA models are suitable especially when model parameters that determine the disease dynamics are unavailable or undetermined due to the disease novelty. In addition, ARIMA models are a flexible, empirical method which is able to produce reliable forecast in situations with limited data. This paper demonstrates that, ARIMA models are able to provide reasonable forecasts even with the above mentioned limitations.

## 2. MATERIALS METHODS

### Data source

Data for this study were obtained from March 21^st^, 2020 to November 26^th^, 2020. The daily Algerian COVID-19 confirmed cases were sourced from The Ministry of Health, Population and Hospital Reform. Daily COVID-19 confirmed cases for neighboring countries were also obtained from the Johns Hopkins University’s official website [6].

### Methods

The Exponential Smoothing method and Autoregressive Integrated Moving Average (ARIMA) processes refer to [16] with the following equation:

- Determine the first smoothing value and determine the parameter *α*

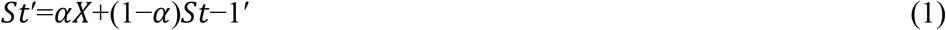
- Determine the second smoothing value

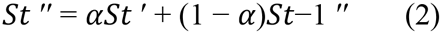
- ARIMA Model For Time Series Data ARIMA model is stated as follows:

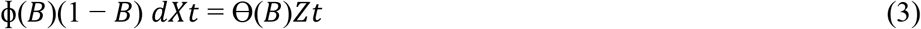

ARIMA forecasts on its previous past values and portrayed by 3 terms-p, d, q. Where, **p** is the order for the Auto Regressive expression (AR), **q** is the order for the Moving Average expression (MA) and **d** is the Number of differencing required making the time arrangement fixed.

Our goal is to that optimizes the metric of interest [17]. The experiment is carried out in Minitab 17 Programming software. In general equation can be approached using a regression model:

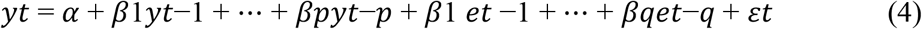

## 3. RESULTS

Using the time-series model approach, the pattern of COVID-19 data distribution behavior in Algeria shows an exponential distribution pattern, where the addition of positive cases of COVID-19 increases significantly every day of the epidemic. This condition is also followed by a distribution pattern of the number of people who recovered and died (**Figure 1**). As we know that in the time-series model the type of exponential distribution consists of a single exponential, double exponential, and Winters’ method. Based on the literature reviews, the best model is the double exponential model [18].

**Figure 1.**
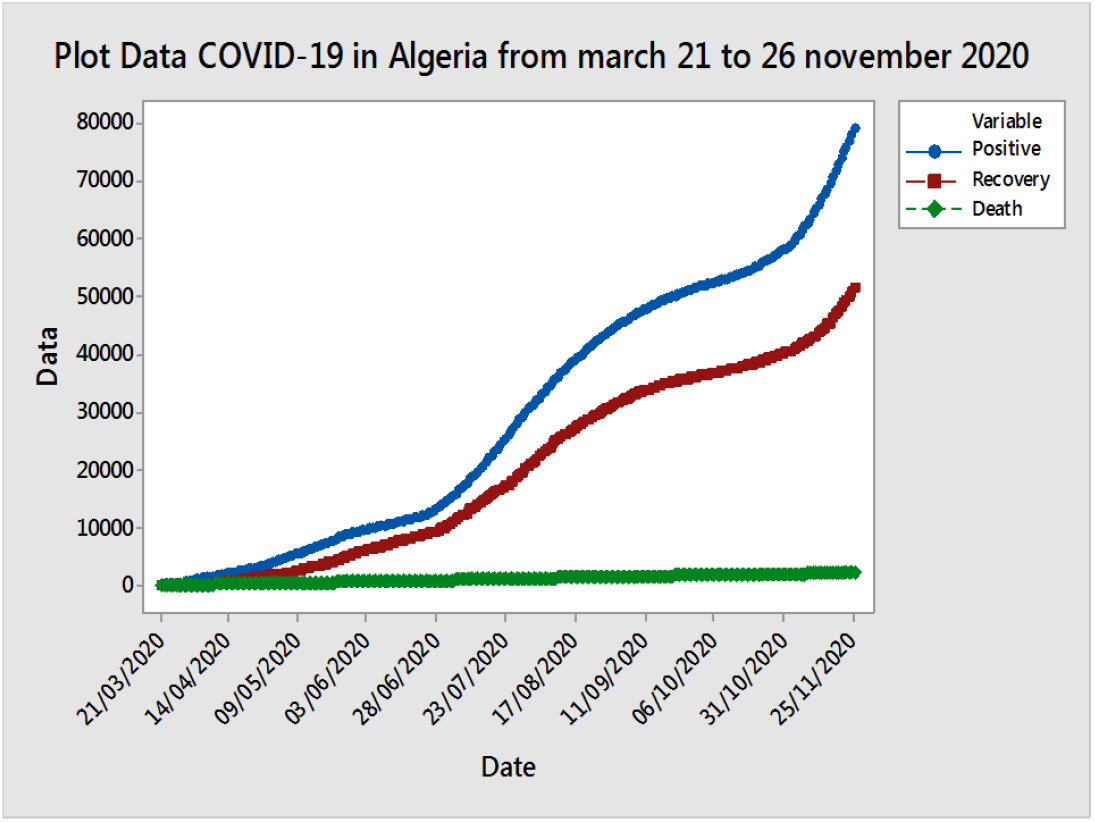
Plot Data COVID-19 in Algeria (Data processed by Minitab 17)

By using a 10 percent error rate, the best α parameter values are 0.745 and the best γ is 1.421 with a Mean Absolute Percentage Error (MAPE) of 0.49 percent. MAPE is the error value for each period divided by the actual observation value for that period (**Figure 2**). In positive COVID-19 cases, the MAPE value is smaller than the error rate at 10%.

**Figure 2.**
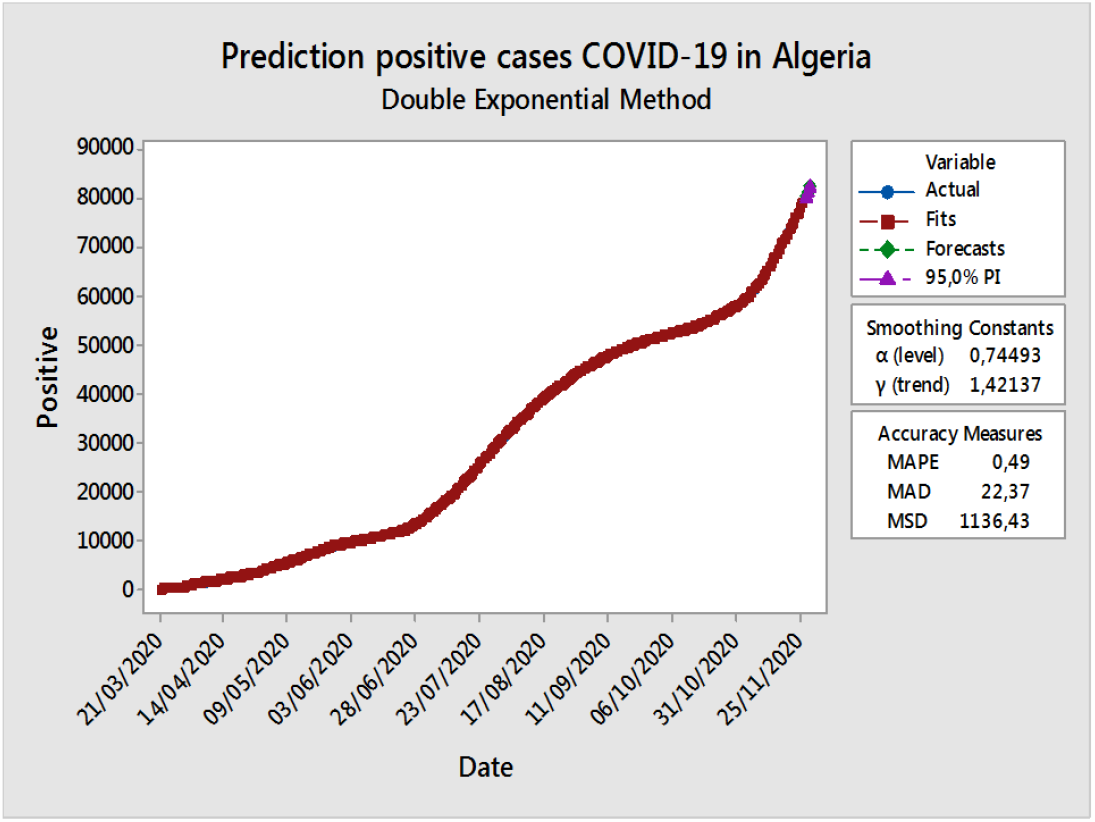
Prediction positive COVID-19 cases in Algeria (Data processed by Minitab 17)

The increase in the number of people who were positive for COVID-19, also directly affected the model of prediction patients who recovery and death **(Figure 3 and Figure 4)**.

**Figure 3.**
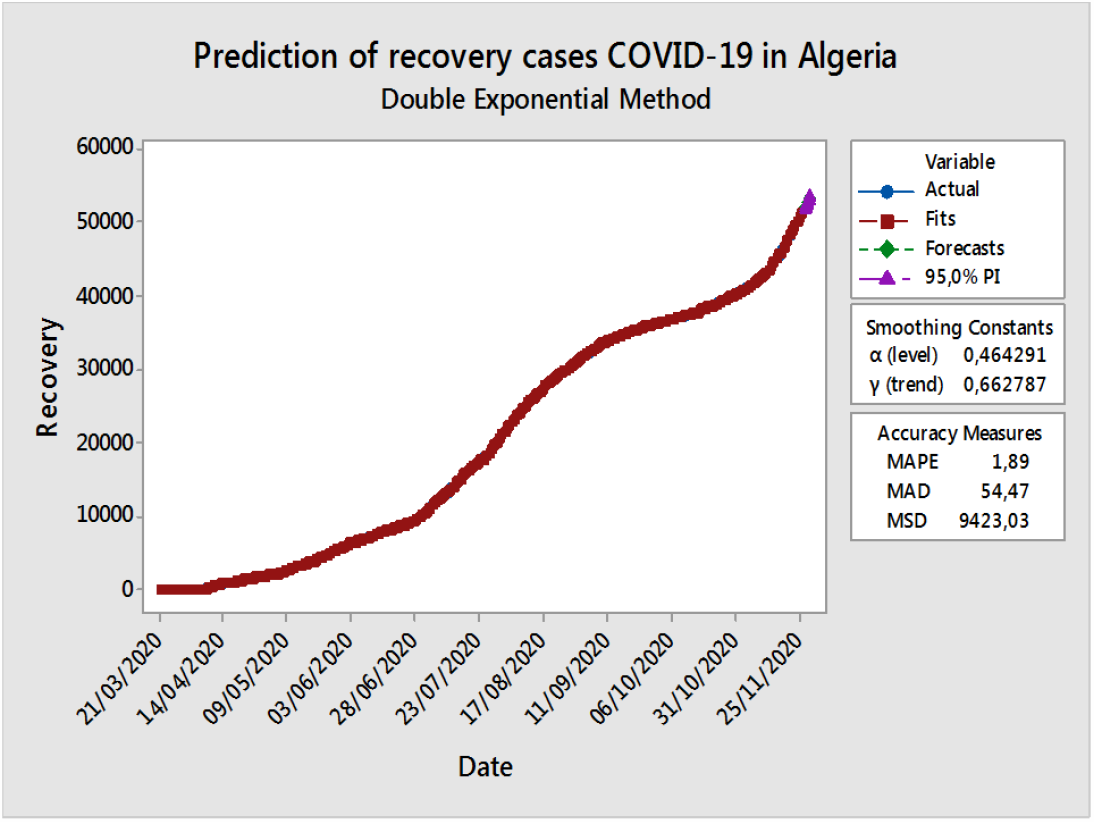
Prediction Recovery COVID-19 cases in Algeria (Data processed by Minitab 17)

**Figure 4.**
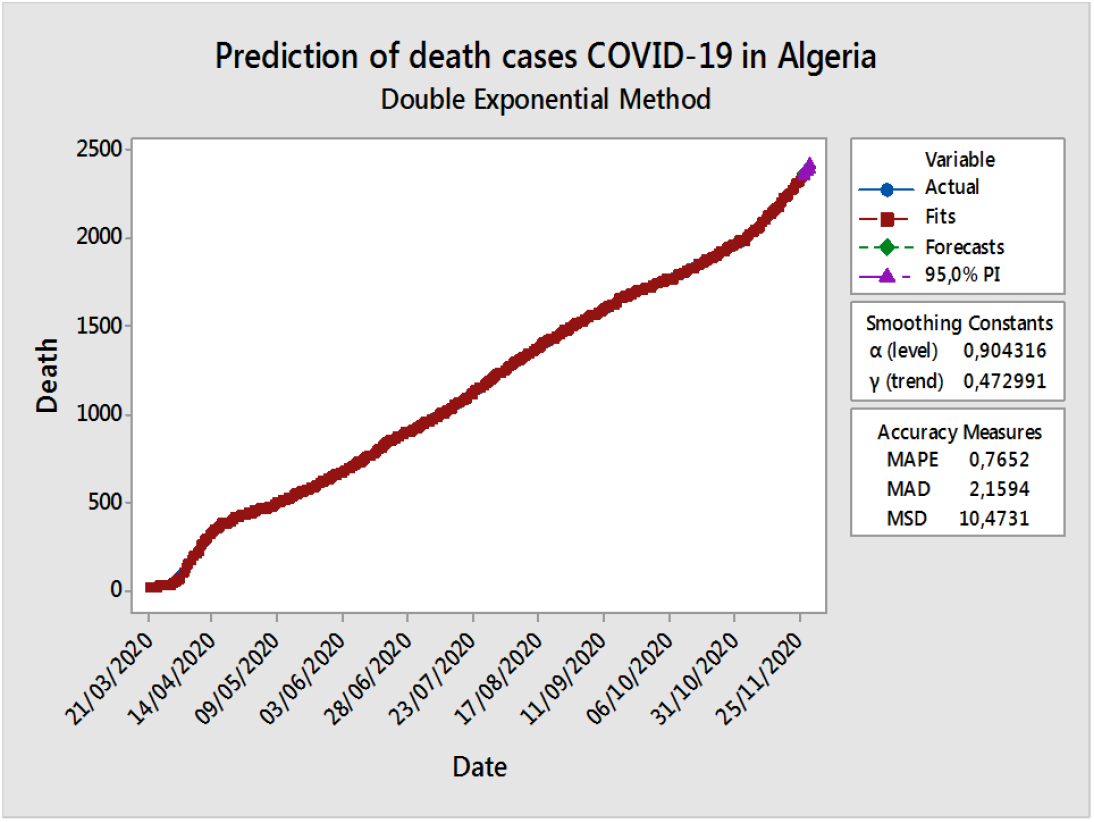
Prediction Death COVID-19 cases in Algeria (Data processed by Minitab 17)

Based on **Figure 3**, using a 10 percent error rate, the estimated value of the parameter recovery patients at α is 0.464, γ is 0.662 and the MAPE of 1.89 percent. In the recovery of COVID-19 cases, the MAPE value is smaller than the error value set at 10 percent error rate. The cure rate of COVID-19 patients is increasing simultaneously with the number of positive cases because of the health measures taken by the government since march 21^st^, 2020, and probably also due to the age of Algerian community which is relatively young.

Using a 10 percent error rate, the estimated value of the parameter death patients at α is 0.904, γ is 0.472, and the MAPE of 0.765 percent. Death cases of COVID-19, the MAPE value is greater than the error value set at 10% (**Figure 4**). The increase in mortality is possibly due to the degree of infection and also the medical history of patients COVID-19.

After the parameter values, α and γ are obtained, the next step is the identification process Partial Autocorrelation Function (PACF) and Autocorrelation Function (ACF) from positive, recovery, and death data. PACF and ACF tests are used to test the accuracy of the results of the double exponential smoothing model and a means of determining the stationarity of the variable and the lag lengths of the ARIMA model.

Based on Figure 5, PACF and ACF plots of Residuals for COVID-19 positive data are obtained. The lag times through PACF cuts off at lag one, and ACF tails off slowly.

**Figure 5.**
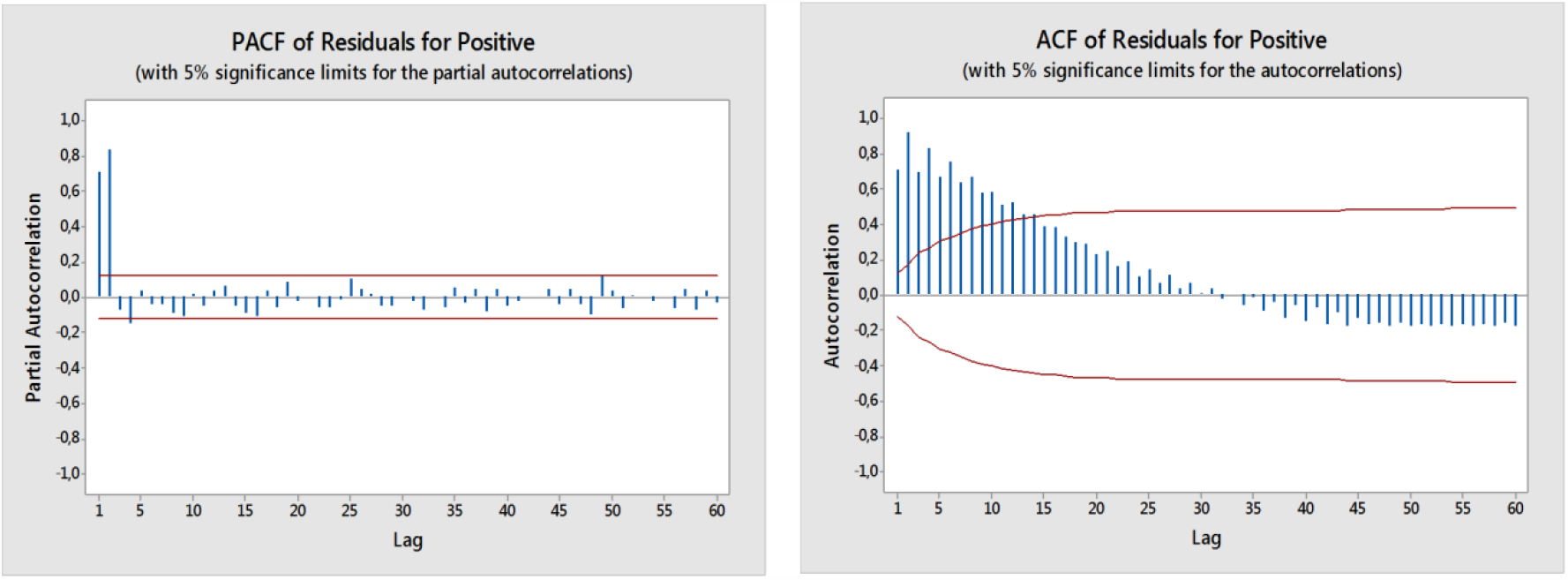
PACF and ACF of Residuals for COVID-19 for Positive Data (Data processed by Minitab 17)

In the time series model with the error probability (α) 5%, the graph follows the ARIMA process (0,1,1) with the p-Value MA 1 (0.0%) is smaller than α. The estimated results of parameters model for COVID-19 Positive Data using ARIMA model are shown in **Table 1**.

**Table 1.**
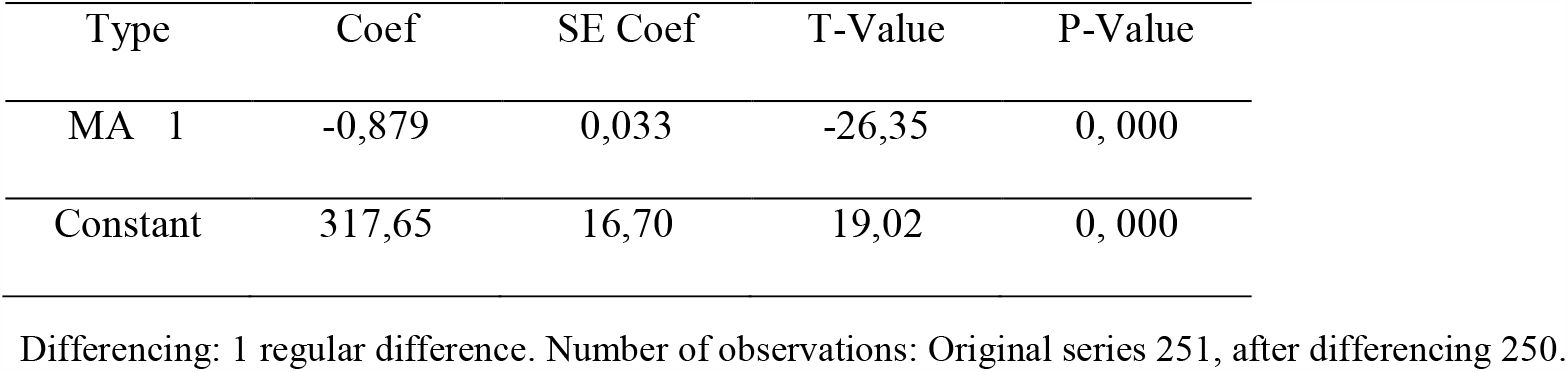
Final Estimates of Parameters Model for COVID-19 Positive Data.

| Type | Coef | SE Coef | T-Value | P-Value |
| --- | --- | --- | --- | --- |
| MA 1 | -0,879 | 0,033 | -26,35 | 0, 000 |
| Constant | 317,65 | 16,70 | 19,02 | 0, 000 |

Differencing: 1 regular difference. Number of observations: Original series 251, after differencing 250.

Referring to equation (4), mathematically the ARIMA model (0,1,1) can be stated using coefficients in **Table 1** as follows: y_*t*_ = 317.65 − 0.879*e*_*t*−1_

With the same steps as testing positive data, next step the identification process PACF and ACF from recovery data.

Same as COVID-19 positive data, **Figure 6** to show PACF and ACF plots of Residuals for COVID-19 Recovery data are obtained. The lag times through PACF cuts off at lag one and ACF tails off slowly. In the time series model with the error probability (α) 5%, the graph follows the ARIMA process (0,1,1) with the P-Value MA 1 (0,0%) is smaller than α. The estimated of parameters for COVID-19 Recovery data results using ARIMA model are reported in **Table 2**.

**Table 2.** Final Estimates of Parameters Model for COVID-19 Recovery Data.

| Type | Coef | SE Coef | T-Value | P-Value |
| --- | --- | --- | --- | --- |
| MA 1 | -0,30 | 0,060 | -4,93 | 0, 000 |
| Constant | 205,53 | 13,14 | 15,64 | 0, 000 |

Differencing: 1 regular difference, Number of observations: Original series 251, after differencing 250.

**Figure 6.**
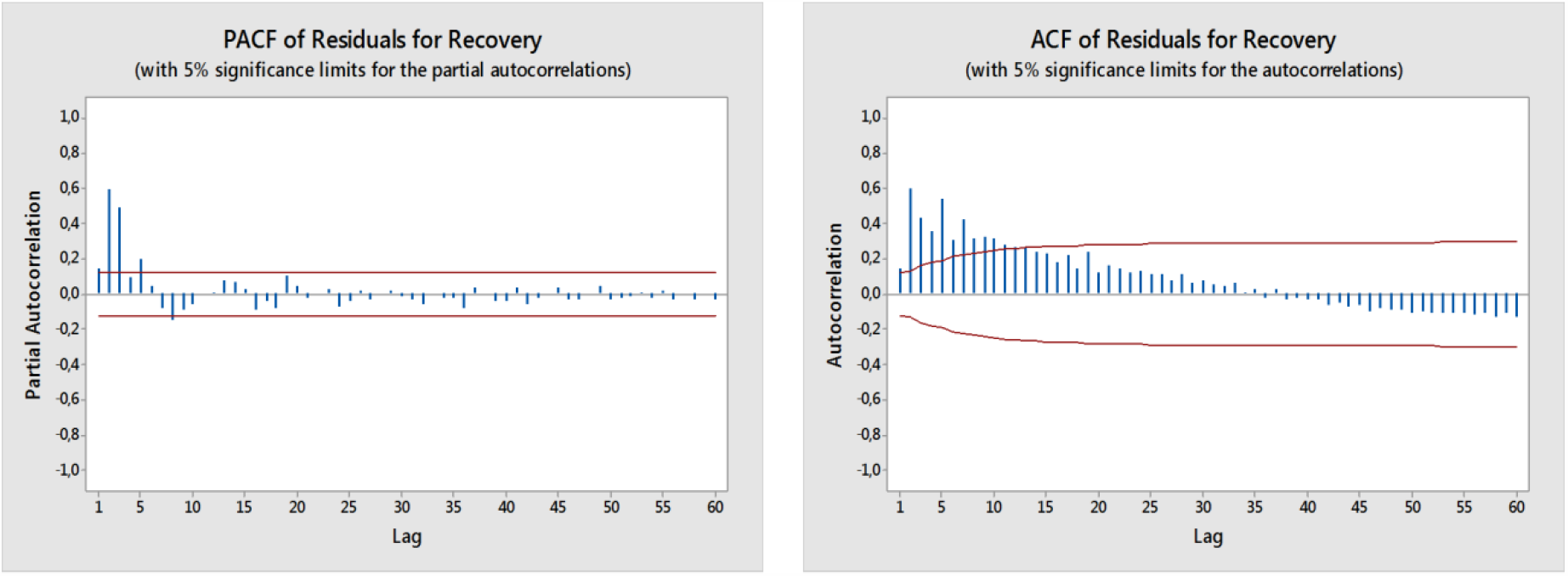
PACF and ACF of Residuals for COVID-19 Recovery Data (Data processed by Minitab 17)

Referring to equation (4), mathematically the ARIMA model (0,1,1) in **Table 2** can be stated as follows: y_*t*_ = 205.53 − 0.30*e*_*t*−1_

After positive and recovery data are analyzed, next the PACF and ACF models of the data death are shown in **Figure 7**. The PACF and ACF plots of residuals for COVID-19 death data are obtained. The lag time through PACF cuts off at lag two and ACF tails off slowly.

**Figure 7.**
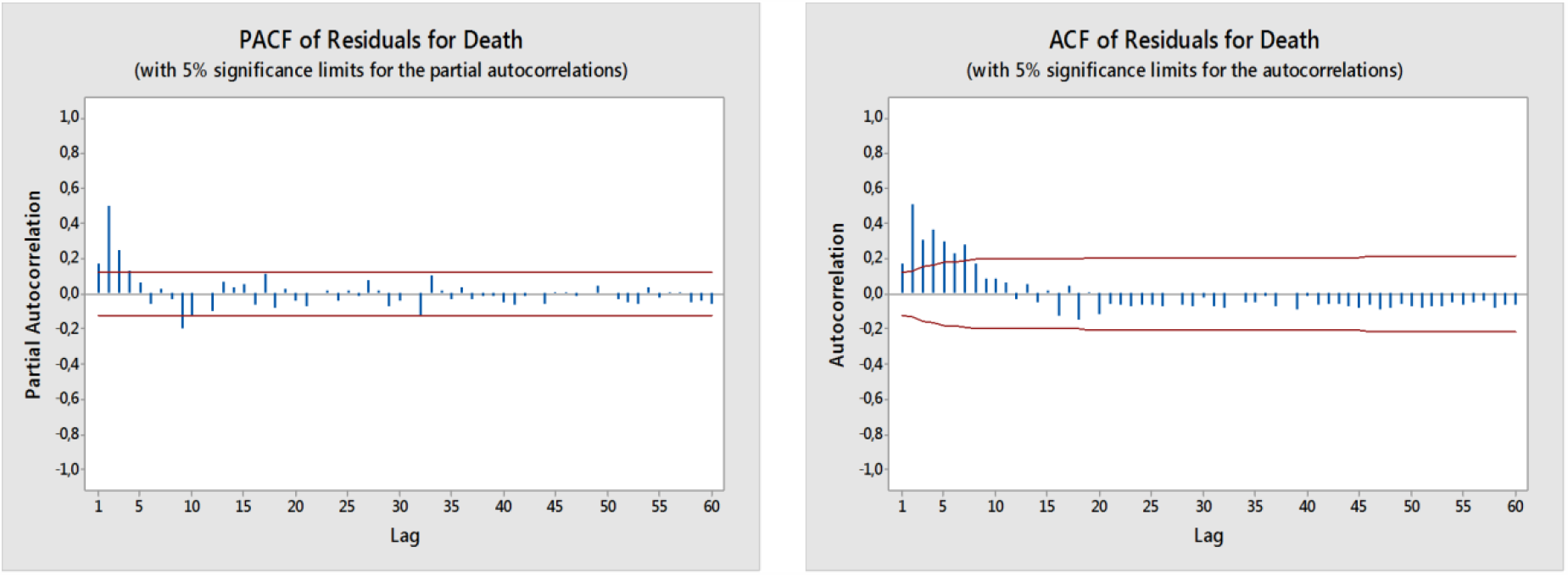
PACF and ACF of Residuals for COVID-19 Death Data (Data processed by Minitab 17)

In time series model with the error probability (α) 5%, the graph follows the ARIMA process (0,1,1) with the p-Value MA 1 (0.00%) is smaller than α. All estimated parameters results of the ARIMA model are shown in the **Table 3**.

**Table 3.** Final Estimates of Parameters Model for COVID-19 Death Data.

| Type | Coef | SE Coef | T-Value | P-Value |
| --- | --- | --- | --- | --- |
| MA 1 | -0,415 | 0,056 | -7,19 | 0, 000 |
| Constant | 9,343 | 0,363 | 25,71 | 0, 000 |

Differencing: 1 regular difference. Number of observations: Original series 251, after differencing 250.

Referring to equation (4), mathematically the ARIMA model (0,1,1) can be stated using constant coefficient’s obtained in **Table 3** as follows: y_*t*_=9.343−0.415*e*_*t*−1_

Based on the results of predictions of COVID-19 cases that occurred in Algeria with a double exponential model and the results of PACF, ACF and estimated parameters model following the ARIMA model (0,1,1) with the p-Value MA 1 is smaller than α. The results of predictions of COVID-19 cases that occurred in Algeria (positive, recovery, and death) showed a gap in the resulting distribution patterns. Where the increase in the number of positive cases has not been offset by an increase in the number of patients who recovered and a decrease in the number of patients who died. This indicates that public behavior still does not comply with the rules set by the government (social distancing, large-scale social restrictions, mask use), the limited medical staff and the lack of standard equipment handling COVID-19 is one of the causes of the low handling of healing from positive patients.

## 4. DISCUSSION

Since the WHO declared COVID-19 a pandemic in March 11^th^, 2020, several countries including Algeria experienced an exponential rise in COVID-19 cases [7]. This rapid increase of cases has stressed most healthcare systems worldwide and has further made outbreak response and resource planning a challenge. In response, health authorities have attempted to forecast the trend of this pandemic, however this have proven to be difficult as COVID-19 is a novel disease with limited data and knowledge on the disease trends and dynamics [6]. This is especially observed when using ARIMA model to predict disease trends, where ARIMA model require sufficient long time series data to be accurate.

Our forecast also showed an accurate trend which corresponded to the positive cases observed and reported by the ministry of health in Algeria during three days (252, 253 and 254). The same situation has been obtained for forecasted recovery and death cases.

As shown in **Table 4**, this finding is strengthened by variations of less than 5% between the forecast and observed cases in 100% of the forecasted data points. This paper demonstrates that ARIMA models are a suitable tool to forecast case trends especially during situations where data is limited.

**Table 4.** Validation of ARIMA model (forecasted positive, recovery and death cases) with 5% significance limits.

| Days forecasted | Forecasts positive cases |  |  |  | Forecasts recovery cases |  |  |  | Forecasts death cases |  |  |  |
| --- | --- | --- | --- | --- | --- | --- | --- | --- | --- | --- | --- | --- |
|  | Forecast | Lower | Upper | Actual | Forecast | Lower | Upper | Actual | Forecast | Lower | Upper | Actual |
| 252 | 80172.9 | 80118.1 | 80227.7 | **80168** | 51958.5 | 51825.1 | 52092.0 | **51946** | 2372.04 | 2366.75 | 2377.33 | **2372** |
| 253 | 81240.6 | 81129.2 | 81352.0 | **81221** | 52579.8 | 52414.9 | 52744.7 | **52568** | 2392.53 | 2384.94 | 2400.11 | **2393** |
| 254 | 82308.3 | 82139.5 | 82477.2 | **82221** | 53201.1 | 53001.9 | 53400.3 | **53204** | 2413.02 | 2402.99 | 2423.04 | **2410** |

Bold values are the reel values reported by the ministry of health and hospital reform of Algeria.

Similarly, studies on COVID-19 conducted in countries such as South Korea, Iran and Italy were able to predict case trends using ARIMA models in similar conditions. In addition as with our findings, a study in Italy also reported a high level of forecast accuracy of 95% in predicting COVID-19 trends using ARIMA models [14-19-20].

The strengths of this study include, firstly, this paper is the first to report the use of ARIMA models to forecast COVID-19 cases and trends in Algeria. Secondly, this was the first attempt to use smoothen case data to improve accuracy as compared to similar studies on ARIMA models for COVID-19 conducted in other countries [20-21]. Thirdly, we used several independent covariates which provided more accurate signals to develop short-term model predictions for immediate outbreak response. And finally, we also optimized the model training and validation period to provide the highest number of data points to generate the best fit model.

## 5. CONCLUSION

This study demonstrated the effectiveness of ARIMA models as an early warning strategy that can provide accurate COVID-19 forecasts on larger data points (251 days). ARIMA models are not only effective but it’s a simple and easy tool by which COVID-19 trends can be predicted based on open access data. The forecasted values of positives, recovery and death cases of COVID-19 shown an accurate trend which corresponded to the actual cases observed and reported by the ministry of health in Algeria during three days (252, 253 and 254). In addition, the use of smoothened data and independent covariates improved the model accuracy. We are confident that the ARIMA model can be used to generate accurate and reliable forecasts of daily COVID-19 cases until the end of COVID-19 with the addition of new data points and independent covariates.

## Data Availability

Conflict of Interest
I like inform you that this is my first submission and this work was never submitted in any participation cited in your ethics section.
For the literature cited in the manuscript, it has been fully acknowledged.
The submission of this paper has been confirmed by all authors cited after my name and followed by their participation to the correction. So, all authors agree with this submission. The authors declare that they have no conflict of interest.
Dr. MESSIS Abdelaziz

## Acknowledgement

The authors would like to thank the Ministry of Health, Population and Hospital Reform of Algeria. We also thank the John Hopkins University for publicly releasing the updated datasets on the number of infected cases of COVID-19.

